# Detection of viruses of public health importance in wastewater samples using conventional PCR techniques and a targeted enrichment whole genome sequencing panel

**DOI:** 10.64898/2026.03.05.26347709

**Authors:** Gonzalo M. Castro, Florencia Mallou, Gisella Masachessi, María C. Frutos, Verónica E. Prez, Tomás Poklepovich, Silvia V. Nates, María B. Pisano, Viviana E. Ré

## Abstract

Wastewater-based epidemiology (WBE) is an effective surveillance approach for monitoring viruses of public health relevance at the community level, complementing clinical surveillance systems. Molecular methods such as PCR/qPCR are widely used for targeted detection, while next-generation sequencing (NGS) with targeted enrichment panels has emerged as a complementary strategy for broader viral detection and genomic characterization. This study comparatively evaluated conventional PCR/qPCR and a targeted enrichment whole-genome sequencing Viral Surveillance Panel (VSP, Illumina) for virus detection in wastewater.

Fifty-six wastewater samples collected between 2017 and 2023 from a wastewater treatment plant in Córdoba, Argentina, were concentrated by polyethylene glycol precipitation and pooled by season and year, reaching a total of 14 pools. Each pool was analyzed in parallel by PCR/qPCR for eight human viruses of public health importance and by the VSP, targeting 66 viral species, sequenced on a NovaSeq 6000 platform, and analyzed with the DRAGEN pipeline.

Detection frequencies for each virus using PCR/qPCR and VSP were: RoV A 100%/14.3%; NoV 100%/14.3%; AiV 50%/42.9%; SARS-CoV-2 14.3%/0%; HAV 42.9%/0%; HEV 14.3%/0%; JCPyV 35.7%/85.7%; BKPyV 28.6%/71.4%, respectively. In addition, VSP detected the genomes of Astrovirus (71.4%), Salivirus (21.4%), Coxsackie A (14.3%), Rotavirus C (14.3%), and Merkel Cell virus (7.1%), and enable the recovery of 16 near complete genomes (coverage > 92.5%) of AiV, JCPyV, BKPyV, Salivirus and Astrovirus.

PCR/qPCR and targeted enrichment NGS provide complementary information wastewater viral surveillance. Their combined application improves virus detection and genomic characterization, reinforcing the value of integrated approaches in environmental virology and public health monitoring.

## Introduction

Wastewater-based epidemiology (WBE) is a powerful tool for the surveillance of microorganisms of public health importance. It provides real-time information on population health by analyzing pathogens excreted in human fluids and feces entering the sewage system (Armas et al., 2023; Masachessi et al., 2024), offering data on the prevalence of pathogens at the community level, complementing clinical surveillance, which is often limited by resources availability, notification bias, and healthcare services access (Girón-Guzmán et al., 2024).

WBE is a fundamental early warning strategy for viral surveillance. Many infected individuals shed viruses before developing symptoms (presymptomatic), or even if they are asymptomatic, allowing incidence peaks or new variants to be detected earlier than with traditional epidemiological strategies, thereby enabling the silent transmission monitoring of these pathogens (Cianella et al., 2023).

The WBE workflow for virus detection includes a) sampling, either random samples taken at a specific time point only once, or composite samples, which involve the collection and pooling of multiple samples over a specific period of time (Cha et al., 2023; Mejías-Molina et al., 2023); b) viral concentration, a critical step due to the low viral load and matrix complexity (Haramoto et al., 2018); c) nucleic acid extraction, using commercial column-based kits or precipitation methods such as the modified TRIzol-based method (O’Brien et al., 2021; Reyes-Calderón et al., 2023); and d) molecular detection and genomic characterization.

Traditionally, polymerase chain reaction (PCR)-based techniques, primarily real-time PCR (qPCR) have been used as reference methods for virus detection and identification in wastewater samples (Polo et al., 2020). However, these techniques can be affected by the presence of inhibitors in the wastewater matrix, which can impact amplification efficiency and subsequent detection and quantification (WHO, 2023).

Next-generation sequencing (NGS) has recently been incorporated into the group of methodologies used for genomic surveillance. It has been reported that this tool allows for more in-depth characterization, monitoring diversity, evolution, and reporting the introduction of new strains (John et al., 2024; Costa et al., 2025). Targeted sequencing using enrichment panels is a strategy that improves viral detection in complex samples such as wastewater. The use of capture probes to specifically enrich viral genomes of interest significantly increases sensitivity, especially in complex samples with low viral loads, facilitating the collection of near-complete genomes (Calabria de Araujo et al., 2024; Cancela et al., 2025).

In either case, due to the complexity of the matrix, these methods require optimization and validation stages prior to their application in WBE (Robins et al., 2022; Paracchini et al., 2024). The objective of this work was to detect viruses in wastewater using traditional molecular techniques (PCR and/or qPCR) and a targeted enrichment whole genome sequencing panel by NGS, and to compare the results obtained.

## Materials and Methods

Fifty-six wastewater samples collected between 2017 and 2023 from the main sewer of the Bajo Grande wastewater treatment plant in Córdoba city (31.4020° S, 64.1063° W), in central Argentina, were selected. The samples were previously concentrated by precipitation with PEG6000 according to the protocol described by Masachessi et al., (2022). Fourteen pools, containing 4 samples each one, were assembled, taking 300 µL of the individual sample: two pools per year, corresponding one to the summer (December, January, February, and March) and one to the winter (June, July, August, and September) of the seven years studied (Fig. 1).

**Fig. 1.**
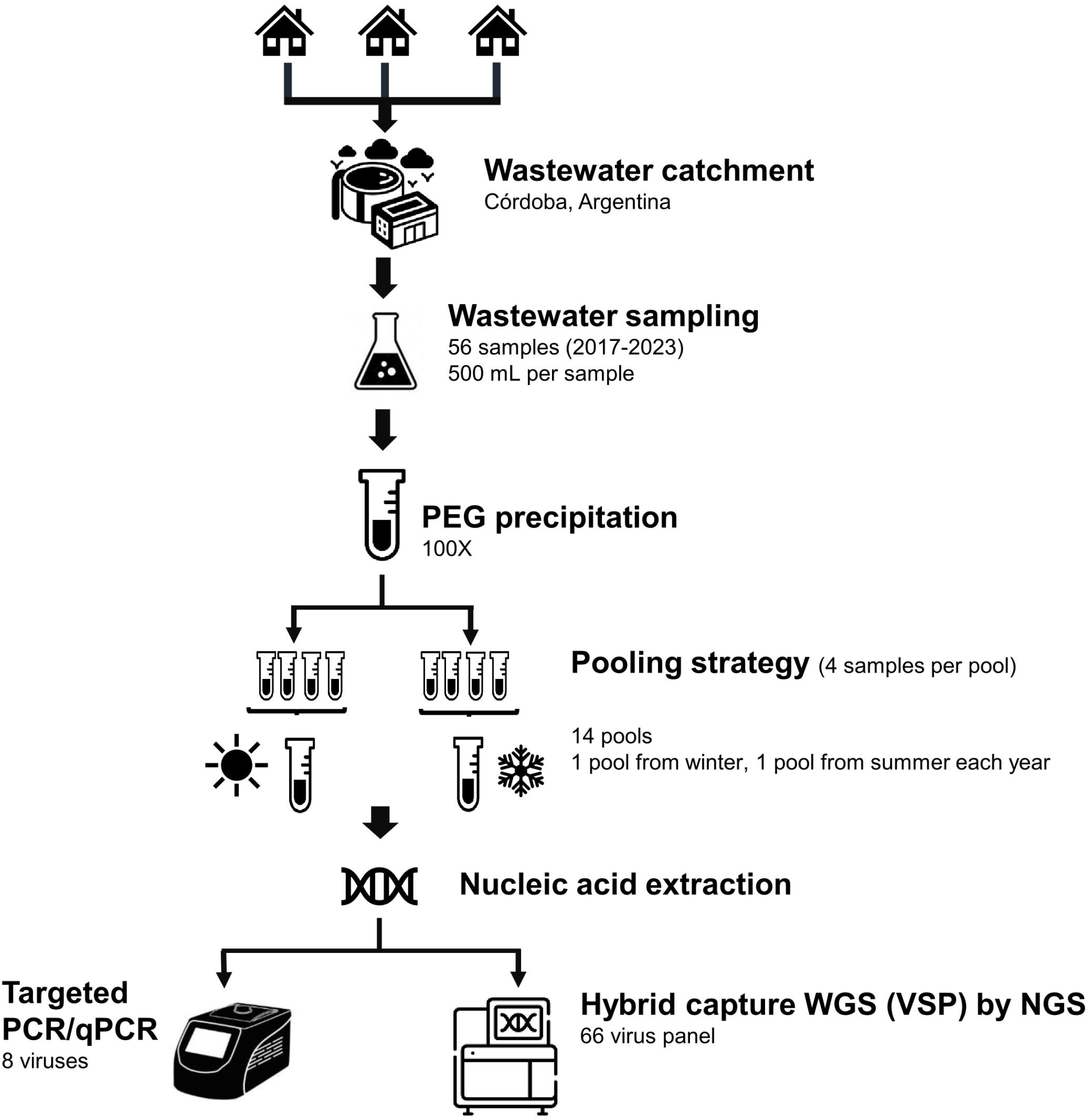
Workflow for virus detection and characterization in wastewater samples by RT-PCR/real time RT-PCR and hybrid capture whole genome sequencing (WGS) using the viral surveillance panel (VSP).

Each pool was subjected to nucleic acid (RNA/DNA) extraction using the MagNA Pure 96 DNA and Viral NA Large Volume Kit - Roche (Mannheim, Germany), following the manufacturer’s instructions. Samples were then processed by PCR/qPCR for the individual detection of 8 viruses: rotavirus (RoV) (Zeng et el., 2008), norovirus (NoV) (ISO 15216-2), Aikivirus (AiV), SARS-CoV-2, hepatitis A virus (HAV), hepatitis E virus (HEV), JC polyomavirus (JCPyV) and BK polyomavirus (BKPyV), according to the protocols described previously (Ré, 2025).

In addition, sample pools underwent genomic surveillance using the Viral Surveillance Panel (VSP) - Illumina (San Diego, CA - USA), following the manufacturer’s instructions. The VSP performs the enrichment of target viral sequences utilizing hybrid capture probes, allowing the detection of complete genomes of 66 viruses (203 strains), including DNA and RNA viruses, identified as high risk to public health. Sequencing was performed on a SP cell, pair-end reads (2×150 bp), assigning 4 million reads to each sample, on a NovaSeq 6000 - Illumina (San Diego, CA - USA). The fastq obtained were analyzed using the DRAGEN Targeted Microbial application available on BaseSpace™Sequence Hub (Illumina). According to the manufacturer’s description, it is expected that of the total reads obtained for each sample: less than 5% will be eliminated due to poor quality, between 30 - 40% will remain unmapped and the rest will correspond to the specific viral sequences detected by the panel.

Nucleotide sequences obtained in this work were deposited in the GenBank database under the accession numbers: PX312153-57 for JCPyV, PX312158-62 for BKPyV and PX390162-65 for AiV.

## Results

Results obtained by PCR or qPCR and VSP for each virus are shown in Table 1. Detection frequencies for each virus using PCR or qPCR/VSP varied substantially across the methods, and were: RoV group A 100%/14.3%; NoV 100%/14.3%; AiV 50%/42.9%; SARS-CoV-2 14.3%/0%; HAV 42.9%/0%; HEV 14.3%/0%; JCPyV 35.7%/85.7%; BKPyV 28.6%/71.4%, respectively. In addition, VSP detected the genomes of Astrovirus (71.4%), Salivirus (21.4%), Coxsackie A (14.3%), Rotavirus group C (14.3%), and Merkel Cell Polyomavirus (7.1%) (Supplementary Table), from which PCR/qPCR were not performed. VSP was able to generate 16 genomes with coverage > 92.5%: 2 of AiV, 5 of JCPyV, 5 BKPyV, 2 of Salivirus and 2 of Astrovirus (Table 1 and Supplementary Table).

**Table 1.** Results of virus detection using PCR/qPCR and the VSP, sequenced on the NovaSeq 6000 system. Coverages corresponding to complete genomes are highlighted in bold.

| Virus | RoV A |  |  | NoV* |  |  | AiV |  |  | SARS-CoV-2 |  |  | HAV |  |  | HEV |  |  | JCPyV |  |  | BKPyV |  |  |
| --- | --- | --- | --- | --- | --- | --- | --- | --- | --- | --- | --- | --- | --- | --- | --- | --- | --- | --- | --- | --- | --- | --- | --- | --- |
| Sample | qPCR (Ct) | Reads | Coverage (%) | qPCR (Ct) | Reads | Coverage (%) | PCR | Reads | Coverage (%) | qPCR (Ct) | Reads | Coverage (%) | qPCR (Ct) | Reads | Coverage (%) | qPCR (Ct) | Reads | Coverage (%) | PCR | Reads | Coverage (%) | PCR | Reads | Coverage (%) |
| Pool 1 | POS (31.9) | nd | nd | POS (30.1) | nd | nd | NEG | 1891 | 81.8 | NEG | nd | nd | NEG | nd | nd | NEG | nd | nd | POS | 716 | 58.5 | POS | 2244 | 97.7 |
| Pool 2 | POS (26.0) | nd | nd | POS (30.3) | nd | nd | POS | nd | nd | NEG | nd | nd | POS (32.8) | nd | nd | NEG | nd | nd | POS | 1453 | 86.3 | NEG | nd | nd |
| Pool 3 | POS (28.0) | nd | nd | POS (28.0) | nd | nd | NEG | 505 | 9.6 | NEG | nd | nd | POS (31.0) | nd | nd | NEG | nd | nd | POS | 2704 | 97.5 | POS | 924 | 68.0 |
| Pool 4 | POS (27.7) | 183 | 12.8 | POS (28.3) | 572 | 25.0 | POS | nd | nd | NEG | nd | nd | POS (33.8) | nd | nd | POS (34.8) | nd | nd | POS | 3100 | 96.3 | POS | 3809 | 97.8 |
| Pool 5 | POS (29.0) | 67 | 7.3 | POS (28.0) | 699 | 13.5 | POS | 6673 | 94.4 | NEG | nd | nd | POS (28.4) | nd | nd | NEG | nd | nd | POS | 5503 | 97.9 | POS | 2707 | 95.0 |
| Pool 6 | POS (28.6) | nd | nd | POS (28.1) | nd | nd | POS | nd | nd | NEG | nd | nd | NEG | nd | nd | NEG | nd | nd | NEG | 1708 | 95.8 | NEG | 1142 | 92.6 |
| Pool 7 | POS (30.9) | nd | nd | POS (31.1) | nd | nd | POS | 10057 | 95.3 | NEG | nd | nd | NEG | nd | nd | NEG | nd | nd | NEG | 3278 | 97.9 | NEG | 389 | 6.1 |
| Pool 8 | POS (29.2) | nd | nd | POS (31.3) | nd | nd | NEG | nd | nd | NEG | nd | nd | NEG | nd | nd | NEG | nd | nd | NEG | 569 | 55.4 | NEG | 1329 | 93.0 |
| Pool 9 | POS (31.3) | nd | nd | POS (28.1) | nd | nd | POS | 222 | 5.9 | NEG | nd | nd | NEG | nd | nd | NEG | nd | nd | NEG | 652 | 66.1 | NEG | nd | nd |
| Pool 10 | POS (27.2) | nd | nd | POS (29.3) | nd | nd | NEG | nd | nd | NEG | nd | nd | NEG | nd | nd | NEG | nd | nd | NEG | 574 | 57.2 | NEG | 311 | 8.0 |
| Pool 11 | POS (29.4) | nd | nd | POS (26.3) | nd | nd | POS | LC | - | POS (27.0)** | nd | nd | POS (36.6) | nd | nd | POS (38.0) | nd | nd | NEG | nd | nd | NEG | LC | - |
| Pool 12 | POS (24.3) | nd | nd | POS (25.9) | nd | nd | NA | nd | nd | NEG | nd | nd | NEG | nd | nd | NEG | nd | nd | NEG | 240 | 5.6 | NEG | 463 | 37.1 |
| Pool 13 | POS (28.9) | nd | nd | POS (26.9) | nd | nd | NEG | 1053 | 60.8 | NEG | nd | nd | POS (36.7) | nd | nd | NEG | nd | nd | NA | LC | - | NA | nd | nd |
| Pool 14 | POS (27.6) | nd | nd | POS (26.2) | nd | nd | NEG | nd | nd | POS (28.8)** | nd | nd | NEG | nd | nd | NEG | nd | nd | NA | 387 | 21.9 | NA | 441 | 41.5 |

nd: no detection
NA: Not Available
LC: low Confidence
\* GII.P7 - GII.6 / GII.P16 - GII.2
ORF1ab

Mean sequencing depth of the obtained complete genomes was: 59X and 109X for AiV; 26X, 32X, 34X, 40X and 60X for JCPyV; 20X, 26X, 30X, 32X and 45X for BKPyV; 43X and 115X for Salivirus and 31X and 181X for Astrovirus.

On average, only 0.4% (range: 0.1 - 1.2 %) of the obtained reads corresponded to specific data that was mapped against a reference genome of the agents detected in the VSP, 2.2% (range: 1.6 - 2.8 %) were eliminated due to poor quality, and 97.5% (range: 97.0 - 98.0%) remained unmapped.

## Discussion

Wastewater viral surveillance has become established as a valuable complementary epidemiological tool for population-based pathogen monitoring, enabling their early detection and community follow-up, especially in contrast to traditional systems based on clinical cases, which often suffer from reporting delays, underdiagnosis, and limited coverage (Prado et al., 2023). In this context, molecular methodologies become relevant, and PCR and qPCR are the ones that have been chosen due to their easy accessibility. In recent years, whole-genome sequencing (WGS) techniques using NGS technology have been added, which have also proven to be effective in the detection and characterization of viruses from aqueous samples (Child et al., 2023).

In this study, some viruses were detected only by PCR/qPCR and not by the VSP panel, indicating a difference in sensitivity between the methods. PCR/qPCR are notable for their high sensitivity and specificity, being efficient for viral screening. These molecular techniques are relatively rapid methods widely available in public health laboratories, making them ideal for routine and cost-effective detection. In fact, many in-house and commercial PCR/qPCR protocols for clinical samples have shown exceptionally good performance in wastewater monitoring for certain viruses (Ré, 2025), even demonstrating a higher detection rate compared to WGS in some cases (Calabria de Araujo et al., 2024; Williams et al., 2024). In our study the overall detection frequency using PCR/qPCR was 1.7 times higher than VSP, in line with what was previously reported.

In our study, although PCR and qPCR methods have proven to be more sensitive than VSP, they are limited to monitoring one or only a few specific target viruses per assay. Furthermore, their performance in complex environmental matrices such as wastewater, can be affected by the presence of inhibitors, RNA degradation, and genetic variability, highlighting the need to adapt and optimize protocols for these non-conventional matrices (de Araujo et al., 2025).

In contrast, hybrid capture approaches can offer an alternative strategy for monitoring a broad spectrum of viruses in wastewater, including their detection and genetic characterization (Martínez-Puchol et al., 2020; Child et al., 2024). In our case, the VSP used, allows for the detection of a wide range of target viruses, including novel and emerging pathogens, and, theoretically, provides uniform coverage of complete viral genomes. This technique reduces unnecessary sequencing of host and non-target microbes compared to shotgun metagenomic sequencing, where the entire RNA/DNA is sequenced, which in turn reduces costs and allows for the comprehensive sequencing of viral genomes (Calabria de Araujo et al., 2024; de Araujo et al., 2025). Furthermore, the oligonucleotide probes used in the VSP kit were designed to maintain their effectiveness in mutated regions, which is crucial for capturing rapidly evolving viruses, such as RNA viruses. Despite all these advantages described, hybrid capture approaches also involve significantly higher costs, advanced infrastructure requirements, and longer processing times, which limits their routine application.

Although all described, during this study we obtained a high number of reads that did not specifically map to any of the VSP targets (97.5%), which could be due to the complexity of the wastewater sample matrix, containing a high bacterial load, human DNA and interfering molecules (such as detergents, metals and humic acids) (de Araujo et al., 2025). This would affect the specificity of the probes used for viral enrichment, making it difficult to detect the viruses present in the samples (Fernández-Cassi et al., 2018; Martínez-Puchol et al., 2020; Child et al., 2024).

In line with the above, it is striking that HAV, HEV, and SARS-CoV-2 were not detected by the VSP in any of the analyzed pools, even though they presented positive results by qPCR in the same years. Indeed, the periods when wastewater samples were collected coincided with documented outbreaks of hepatitis A and with the COVID-19 pandemic (Mariojoules et al., 2023; Masachessi et al., 2022; Fantilli et al., 2023; Fantilli et al., 2024; Masachessi et al., 2024). Furthermore, gastroenteritis-associated viruses such as rotavirus (RoV) and norovirus (NoV), which show high circulation in our setting (ANLIS - Malbrán 2022; 2023; Gomes et al., 2025), were consistently detected in all samples by PCR/qPCR but were identified by VSP in only two samples each one.

The wastewater samples used in this study were processed by centrifugation to remove solids before viral enrichment by precipitation with PEG6000. This method has been standardized and validated as a valid protocol for use in enteric virus surveillance by PCR/qPCR, for which it has provided adequate sensitivity (Masachessi et al., 2018; Masachessi et al., 2022; Fantilli et al., 2023; Fantilli et al., 2024; Masachessi et al., 2024). However, its efficiency for virus detection using VSP remains unknown and may partially explain some of the discrepancies observed in our results, such as the ones mentioned above (the non-amplification of HAV, HEV or SARS-CoV-2), as well as the low coverage depths obtained. Nevertheless, the proportion of viral reads recovered in this study was comparable to that reported in other wastewater studies employing VSP and using other concentration methods (Calabria de Araujo et al., 2024; de Araujo et al., 2025), which suggests that additional factors may be involved in the interference and non-detection of certain viruses.

In this regard, the pre-analytical treatments currently used were developed in the pre-NGS era, without considering certain factors that could influence viral detection by this type of technology. Therefore, there is a need to reassess pre-analytical workflows, particularly to address the high abundance of non-viral DNA/RNA that may hinder the amplification and detection of viruses present at low concentrations in environmental samples. Implementing new wastewater pre-analytical processing strategies has become a key technical challenge for improving the performance of viral genomic detection. DNase or RNase pretreatment to remove extracellular and extra viral nucleic acids before extraction could be an option, in order to enrich the viral nucleic acid fraction. However, this treatment could have an undesirable side effect, eliminating the nucleic acid of lysed enveloped viruses due to the detergents present in the matrix, so its use should be investigated before being implemented in this type of sample (Ye et al., 2016; Schuele et al., 2021). These considerations and recommendations should be considered and incorporated into the inserts and protocols of virus genomic detection panels using NGS.

According to our records, this is the first study in Argentina to use hybrid capture probes for the detection and genetic characterization of multiple viruses in wastewater, which allowed to obtain the first complete genomes of poorly studied viruses such us AiV, polyomavirus and Salivirus.

Taking into account all the results, the combined application of PCR/qPCR and VSP methodologies in wastewater viral surveillance offers the most practical and effective balance. While PCR provides the sensitivity and speed necessary for viral screening of the main human pathogens of public health concern, the VSP provides valuable genomic information on these and other understudied viruses, offering the genomic depth and broad-spectrum detection capacity necessary for molecular epidemiology. This combination of methodologies has become a key tool for virus surveillance that could help public health authorities to guide preventive and control measures.

## Data Availability

All data produced in the present work are contained in the manuscript

## Notes

### Competing Interest Statement

The authors have declared no competing interest.

### Funding Statement

This work was supported by grants PICT 2021 Cat II 00041 (Foncyt) and CB 2 PFI2022 Res 2022 725 APN MCT (Cofecyt), from the National Government of Argentina.

