## Supplementary Table for "Detection of viruses of public health importance in wastewater samples using conventional PCR techniques and a targeted enrichment whole genome sequencing panel"

**Supplementary table:** Results of virus detection using the VSP, sequenced on the NovaSeq 6000 system. Coverages corresponding to complete genomes are highlighted in bold.

| **Virus** | **Astrovirus** | | **Salivirus** | | **Coxsackie A** | | **RoV C** | | **Merkell Cell**  **Polyomavirus** | |
| --- | --- | --- | --- | --- | --- | --- | --- | --- | --- | --- |
| **Sample** | Reads | Coverage (%) | Reads | Coverage (%) | Reads | Coverage (%) | Reads | Coverage (%) | Reads | Coverage (%) |
| **Pool 1** | nd | nd | 4629 | **98,6** | 399 | 32,1 | nd | nd | nd | nd |
| **Pool 2** | 413 | 12,4 | nd | nd | nd | nd | 2475 | 65,0 | nd | nd |
| **Pool 3** | 1669 | 87,9 | nd | nd | nd | nd | nd | nd | nd | nd |
| **Pool 4** | 15808 | **97,8** | nd | nd | nd | nd | nd | nd | nd | nd |
| **Pool 5** | 1839 | 69,1 | 14724 | **98,9** | 559 | 25,9 | nd | nd | 343 | 9,4 |
| **Pool 6** | 1755 | **93,6** | nd | nd | nd | nd | nd | nd | nd | nd |
| **Pool 7** | nd | nd | 1117 | 50,1 | nd | nd | LC | - | nd | nd |
| **Pool 8** | nd | nd | nd | nd | nd | nd | nd | nd | nd | nd |
| **Pool 9** | 366 | 16,7 | nd | nd | nd | nd | nd | nd | nd | nd |
| **Pool 10** | 1097 | 72,9 | nd | nd | nd | nd | nd | nd | nd | nd |
| **Pool 11** | LC | - | nd | nd | nd | nd | nd | nd | nd | nd |
| **Pool 12** | 389 | 23,7 | nd | nd | nd | nd | nd | nd | nd | nd |
| **Pool 13** | nd | nd | nd | nd | nd | nd | nd | nd | nd | nd |
| **Pool 14** | 835 | 67,0 | nd | nd | nd | nd | nd | nd | nd | nd |
| **nd: no detection** | |  |  |  |  |  |  |  |  |  |
| **LC: Low Confidence** | |  |  |  |  |  |  |  |  |  |
